# Aging and Postural Changes Influence the Agreement Between Pulse Rate Variability and Heart Rate Variability

**DOI:** 10.1101/2025.09.17.25336015

**Authors:** Runwei Lin, Marjolein Klop, Frank R. Halfwerk, Richard van Wezel, Gozewijn Dirk Laverman, Dirk W. Donker, Ying Wang

## Abstract

**Objective:** Pulse rate variability (PRV) derived from photoplethysmography is often used as a surrogate for heart rate variability (HRV) in daily monitoring. However, their interchangeability in older adults and under postural change remains uncertain, potentially leading to misinterpretation in practical usage. Moreover, the mechanisms underlying HRV–PRV differences are incompletely understood.

**Approach:** We evaluated HRV–PRV interchangeability across 18 features during four kinds of orthostatic tests in 35 younger and 42 older adults. Bland–Altman analysis assessed the interchangeability in different postural change conditions. Linear mixed models (LMMs) quantified the effects of age, sex, and postural changes. Mediation analysis examined whether mean arterial compliance, total peripheral resistance, and cardiac contractility mediated these effects.

**Results:** 16 features were interchangeable at individual level in younger adults, while only seven in older adults. Aging and posture significantly reduced interchangeability. A significant Age×Sex interaction was identified for metrics reflecting slow variation, with older females exhibiting a most pronounced HRV-PRV agreement decline. Mediation analysis showed that while vascular and hemodynamic parameters differed significantly across subgroups, they explained only a limited portion of the age- or posture-related decline in HRV–PRV consistency.

**Significance:** Our findings demonstrate that PRV cannot reliably substitute HRV to capture physiologically fast variability in older adults. The significant age-sex interaction highlights the necessity for calibration models particularly for monitoring elderly female populations. These insights are essential for daily-life monitoring, emphasizing the need for reliable HRV assessment in older adults and after posture change.

## I. Introduction

Heart rate variability (HRV) is a widely used indicator of autonomic and cardiovascular health [1], [2]. Reduced HRV has been linked to various diseases, including myocardial infarction [3], [4], hypertension [5], [6], diabetes [7]–[9], sleep disorders [10], [11], and depression [12]. Electrocardiogram (ECG) based HRV measurement, i.e., the gold standard, usually requires gel electrodes and is not feasible for daily-life monitoring. Photoplethysmography (PPG) offers a practical alternative for ECG using optical approach [13], [14]. PPG-derived HRV is commonly referred to as pulse rate variability (PRV). In recent years, PRV is increasingly used for continuous monitoring in free-living conditions, especially among older adults [15], [16].

Although PRV is widely adopted, it has been suggested that PRV might contain information distinct from HRV, and direct substitution may lead to misinterpretation [13], [14], [17]. While several studies have reported strong HRV–PRV agreement in young, healthy individuals at rest [14], [17], [18], growing literature reported the interchangeability might reduce in people with worse health conditions, and under dynamic situations such as postural change [18], [19]. Evidence from clinical cohorts suggests that PRV cannot substitute HRV in patients with multiple kinds of cardiovascular, pulmonary, and neurological diseases [20]. Observations of HRV-PRV interchangeability during postural changes and exercises are inconsistent: some concluded PRV can substitute HRV [19], [21], [22], whereas others found poor agreement [18], [23]. Aging has also been hypothesized to reduce HRV–PRV inter-changeability by increasing vascular stiffness, thereby altering pulse wave transmission [14], [21]. Notably, it is recognized that there is a sexual difference on vascular aging [24], [25]. Therefore, investigating whether HRV–PRV agreement follows a sex-specific aging trajectory is essential for the use of PRV across diverse demographics. Overall these discrepancies raise concerns about potential misinterpretation when PRV is used as a direct surrogate for HRV in daily monitoring.

The physiological mechanisms underlying HRV–PRV differences are also not fully understood yet. Pulse arrival time (PAT) variability and blood pressure (BP) level have been proposed as the potential contributors. PAT reflects both cardiac electromechanical delay (pre-ejection period) and pulse transit time (PTT), and variability in PAT has been hypothesized to cause the HRV-PRV discrepancies [21], [26]. In recordings from patients in intensive-care-unit, different BP levels have been associated with differences in HRV–PRV agreement, likely due to the close relationship between BP and PAT [27]. However, few studies have quantified to what extent these factors will impact HRV–PRV agreement.

In the context of daily-life health monitoring, aging and postural changes are key considerations of HRV-PRV interchangeability because they profoundly influence cardiovascular regulation and occur universally in everyday activities. Despite the debates and hypotheses in the literature [14], [18], [21], [22], [28], the effects of aging and postural changes on the interchangeability have not been sufficiently investigated so far. Therefore, this study aims to: (1) systematically examine the HRV-PRV agreement in younger and older adults during postural changes. (2) determine the independent contributions of aging, sex, and postural change to HRV–PRV interchangeability and (3) evaluate whether vascular and hemodynamic measures (mean arterial compliance (AC), mean total peripheral resistance (TPR), and cardiac contractility (dP/dt_max_) mediate aging and posture effects.

## II. Materials and methods

### A. Data acquisition and processing

#### 1) Dataset description

This study utilized three datasets. The first dataset on healthy volunteers used in this study was adopted from [29]–[31]. All older adults included were community-dwelling, able to walk at least 250 meters without the use of walking aids and independent for activities of daily living [30]. The original study was approved by the Radboud University ethics committee (ECS17022 and REC18021). As the dataset is anonymized and available open access, no additional ethical approval was required. This dataset was selected as it includes both younger and older adults undergoing well controlled postural changes enabling direct comparisons of HRV and PRV across different age cohorts. Postural change including four conditions: rapid supine-to-stand, slow supine-to-stand, head-up tilting, and sit-to-stand, were done by individual subjects under researchers’ instruction [29], [30]. During the tests, subjects were required to stay in a resting position (sitting or supine) for five minutes and then stood up for three minutes. For the supine-to-stand test, subjects were required to stand up from a supine position in approximately three or 10 seconds for rapid and slow tests, respectively. For head-up tilting, the subjects were passively tilted up to 70-degree in 15 seconds from supine position to upright. In the sit-to-stand test, the subjects were required to stand up from a sitting position at a preferred speed. For further details of the experiments, please refer to [29]–[31].

We categorized the subjects into two groups according to the criteria that age *>* 60 were regarded as older adults and age < 45 were younger adults [32]. As a result, 31 older adults (median age 77, IQR [75, 80.75], 16 females) and 26 younger adults (median age 23, IQR [22, 27], 7 females) were included in the analysis. PPG, ECG and continuous BP were recorded simultaneously with sampling rates of 1kHz, 300Hz, and 200Hz, respectively. PPG was measured from the distal phalanx of the index finger. Five-lead ECG and beat-to-beat BP were recorded by Finapres Nova (Finapres, Enschede, the Netherlands) using a finger cuff and was corrected by a reference height sensor. All signals were synchronized using a common reference analog signal.

The second dataset was collected from the outpatient clinic of geriatric department at Radboud University Medical Center, including 31 adults (70 years or older). The study was approved by the medical ethics committee (CMO Arnhem-Nijmegen, NL70130.091.19) and performed in accordance with the declaration of Helsinki. All subjects signed informed consent. During experiment, they rose from a supine position at their comfortable pace for three repetitions. These maneuvers were thus classified as rapid supine-to-stand test.

Three-lead ECG and continuous BP were acquired at a sampling rate of 200 Hz. Finger PPG was recorded at 1 kHz and downsampled to 200 Hz for synchronization. PPG, ECG and BP were synchronized using pulse block reference signals. Due to missing recordings, 26 subjects were finally included in this study (median age 75, IQR [75, 80], 12 females). For further detail of the dataset, please refer to [33].

As postural transitions frequently introduce significant motion artifacts that affect peak detection, a data segmentation protocol was implemented. For both dataset I and II, the pre-standing segment was defined as a 280-s window, spanning from 290s (10s after lying/sitting) to 10s prior to the posture change. The post-test segment consisted of a 160-s window, spanning from 10s to 170s after the transition to a standing position. The 10s buffer was discarded from the onset and offset of each segment to mitigate residual motion-related contamination. Consequently, each test yielded two segments, with one for each physiological state with no overlap.

The third dataset includes 10 young healthy subjects (median age: 24.5, IQR: [23, 26], 5 females) who performed three repetitions of a 5-minute supine to 5-minute standing. The experiment took place at the eHealth House at University of Twente, the Netherlands. All subjects signed informed consent, and the study was approved by the Ethics Committee Computer and Information Science of the University of Twente (nr. 250290). Due to an incomplete recording in one repetition for one subject, a total of 29 measurements were included in the final analysis. The synchronized ECG measured using a Einthoven triangle (TMSi ECG module) and finger PPG (Nonin 8000AA) were recorded at 4000 Hz using a TMSi SAGA amplifier (Twente Medical System International, Oldenzaal, the Netherlands). Both signal were resampled to 300Hz for analysis. Similarly, these data were categorized into the rapid supine-to-stand test. To ensure temporal alignment with the segmentation protocol above, a 280s pre-standing window and a 160-s post-standing window (10s to 170s post-transition) were utilized for analysis.

#### 2) Peak detection, data selection and preprocessing

Fig. 1 presents an overview of data processing and statistical analysis in this study. R peaks were derived from lead II ECG using Pan-Thompkins method [34]. MSPTDfast was utilized for PPG peak detection given its superior performance [35], [36]. The midpoint of systolic and diastolic peaks is adopted as fiducial points as it gives the best approximation of PRV with reference to HRV [26], [36]. Fig. 2 showed examples of ECG and PPG waveforms with annotated peaks from younger and older adults.

**Fig. 1.**
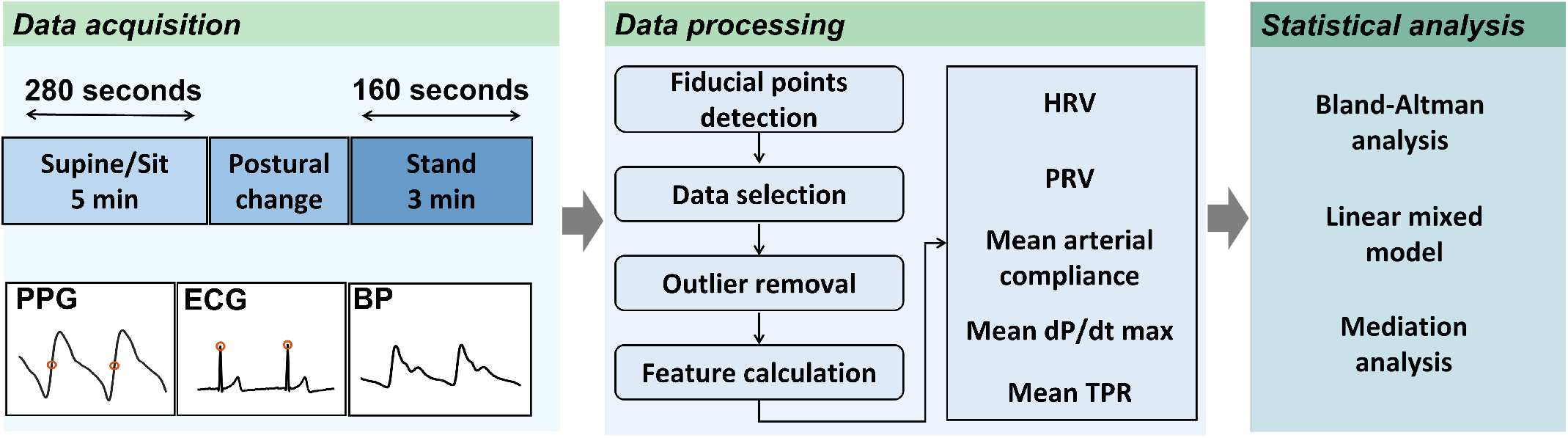
Overview of the study. After fiducial points detection, outlier removal and feature calculation, Bland-Altman analysis were used to assess HRV-PRV agreement. Linear mixed model and mediation analysis were conducted to explore the relevant effects.

**Fig. 2.**
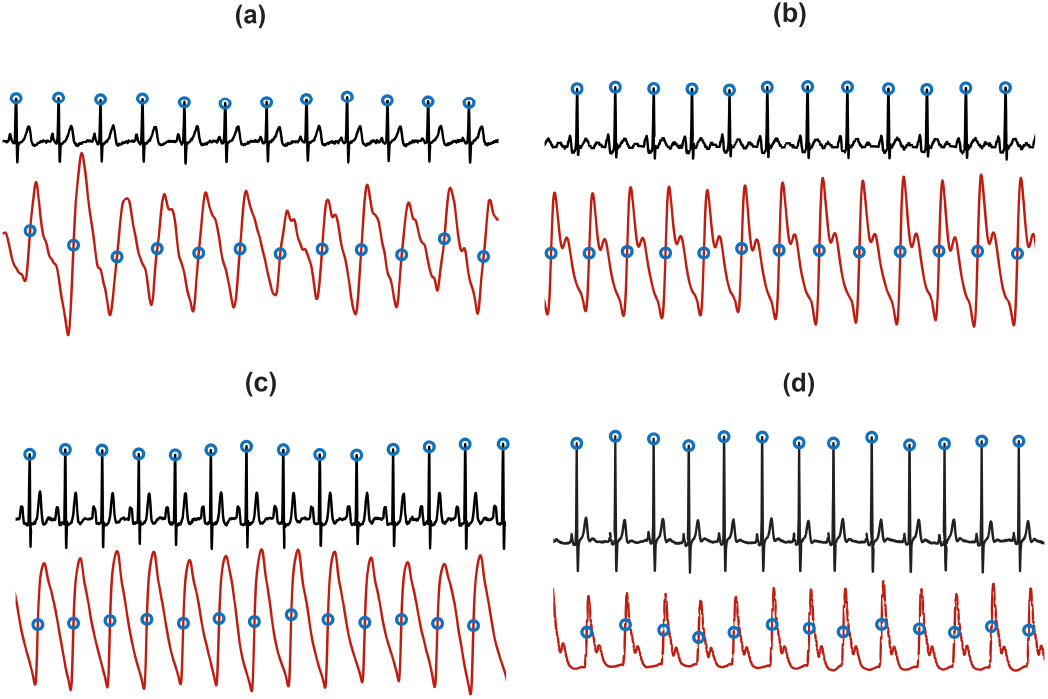
Representative ECG and PPG waveforms with annotated peaks: (a) An older adult from Dataset I; (b) An younger adult from Dataset I; (c) An older adult from Dataset II. (d) A younger adult from Dataset III.

To ensure the reliability of the HRV-PRV consistency analysis, valid segments were selected according to a two-stage quality-control procedure. First, a peak-matching algorithm was implemented to pair detected PPG peaks with their corresponding ECG R-peaks. Within each R-R interval, a valid match was defined based on two constraints: first, interval between the first R-peak and the subsequent PPG peak (pulse arrival time) must fall within 50–500 ms; second, the variation between successive PAT values must be less than 30%. In instances where multiple PPG peaks were detected within a single R-R interval, only the first detected peak was retained. These empirical criteria were established considering the previous literature studying the PAT from different population under resting condition [26], [37]. The relatively broad range was used only to exclude clearly implausible matches while retaining physiologically plausible candidates across subjects. Second, a strict segment-level criterion was applied: only segments with a peak-matching ratio exceeding 95% were retained for analysis. This ensured that the majority of cardiac cycles within each segment had consistent matched peaks. For the subsequent PRV analysis, only these matched PPG fiducial points were utilized to ensure that the pulse intervals directly corresponded to cardiac cycles.

To further explore the mediating role of hemodynamic variables in HRV-PRV consistency difference across subgroups, a secondary subset was derived exclusively from the previously validated segments in Dataset I. Hemodynamic parameters were estimated via a three-element Windkessel model using the software provided by the Finapres (Finapres, Enschede, the Netherlands). To ensure the reliability of the mediation analysis, we retained only recordings where systolic and diastolic blood pressure missingness was below 25%. This relatively large threshold was set empirically, as we only used the mean value of hemodynamic parameters within each segment.

#### 3) HRV and PRV calculation

Prior to calculating the HRV and PRV indices, the resulting inter-beat interval (IBI) series were further refined using a 20% adaptive change rule to eliminate residual ectopic beats or artifacts [38]. Table I summarizes the definitions of the selected features. Since all features were calculated within short time windows, we excluded features with inherently slow variations, such as the very low frequency (≤0.04 Hz) component. In total, 18 features were computed across time, frequency, and nonlinear domains. Time-domain and Poincaré plot features were calculated using the toolbox introduced in [39]. Frequency-domain features were estimated using the Lomb–Scargle periodogram, as recommended in [40], to avoid interpolation-induced errors. Entropy-based features were computed with a tolerance of 20% of the standard deviation and an embedding dimension of m = 2. The detrended fluctuation analysis was performed using the toolbox introduced [41]. The scaling exponents were calculated for two distinct window ranges: 4–16 beats for the short-term exponent (alpha1) and 16–64 beats for the slower exponent (alpha2). To facilitate interpretation, all features were grouped by their physiological timescale into two types: indices reflecting fast and slow physiological regulation fluctuations (Table I).

**Table I.** Summary of definitions, units, and variability types of the selected features.

| Feature [unit] | Definition | Timescale |
| --- | --- | --- |
| AVNN [ms] | Average length of all NN intervals | Slow |
| RMSSD [ms] | Root mean square of the differences between successive NN intervals | Fast |
| SDNN [ms] | Standard deviation of all NN intervals | Slow |
| SDSD [ms] | Standard deviation of the differences between successive NN intervals | Fast |
| pNN50 [%] | Percentage of successive NN intervals differences exceeding 50 ms | Fast |
| LF [$\text{ms}^2$] | Power of low frequency band (0.04-0.15 Hz) | Slow |
| HF [$\text{ms}^2$] | Power of high frequency band (0.15-0.4 Hz) | Fast |
| TP [$\text{ms}^2$] | Total power within 0.04–0.4 Hz | Fast |
| nLF [n.u.] | Low frequency power normalized by TP | Fast |
| nHF [n.u.] | High frequency power normalized by TP | Slow |
| LF/HF | Ratio of LF to HF power | Fast |
| SD1 [ms] | Standard deviation of short axis in Poincaré plot | Fast |
| SD2 [ms] | Standard deviation of long axis in Poincaré plot | Slow |
| SD2/SD1 | Ratio between SD2 and SD1 | Fast |
| ApEn | Approximate entropy of NN intervals | Slow |
| SampEn | Sample entropy of NN intervals | Slow |
| $\alpha_1$ | Faster scaling exponent from detrended fluctuation analysis | Fast |
| $\alpha_2$ | Slower scaling exponent from detrended fluctuation analysis | Slow |

#### B. Bland-Altman analysis

Bland-Altman analysis quantifies whether PRV can replace HRV by evaluating the bias and limit-of-agreement (LoA) between HRV and PRV. The bias refers to the average systematic difference between HRV and PRV, when its 95% CI includes zero, the two measures were considered consistent at the population level, indicating the absence of systematic bias. LoA quantifies the range within which most individual differences fall, with wider LoA indicating poorer agreement. As no universally accepted threshold exists for acceptable limits of agreement in HRV–PRV comparisons, we adopted a predefined operational criterion of LoA width <150% informed by previous studies [18], [42] to indicate acceptable individual-level agreement. When the LoA width exceeded 150%, the two measures were not interchangeable at an individual-level regardless of bias.

These two measures and their 95% CIs were defined as follows. To account for the multiple measurements within each subject, each 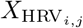 represents the average HRV across all segments for *j*th subject within a specific state (pre- or post-standing). The biases and LoA width were normalized by HRV to facilitate comparisons across different features. Specifically, for pNN50, we calculated the absolute bias and LoA without normalization. This was because many older adults exhibit an ECG-derived pNN50 of 0, which would result in undefined or infinite relative errors during normalization. The result of pNN50 is thus incomparable with other features.

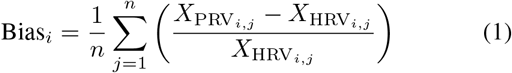

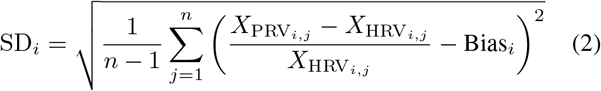

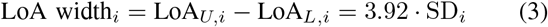

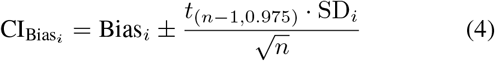

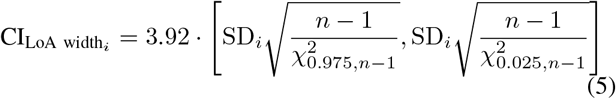

where 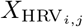 and 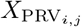 denote the *i*th average HRV and PRV feature for *j*th subject. *n* is the number of subjects within each subgroup. SD_*i*_ is the standard deviation of the normalized HRV-PRV differences, and LoA width_*i*_ is the distance between the upper and lower LoA. LoA_*U,i*_ and LoA_*L,i*_ were distribution upper and lower LoA for *i*th feature. *t*_(*n−*1,0.975)_ is the 0.975 quantile of the *t*-distribution with *n−*1 degrees of freedom, and 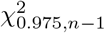 and 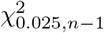 are the corresponding upper and lower quantiles of the chi-square distribution with *n* − 1 degrees of freedom, respectively.

To explore whether PRV can reliably replace HRV under the influence of aging and postural changes, we performed Bland–Altman analysis in four subgroups, denoted by the *Group*–*State* pattern: younger-pre, younger-post, older-pre, and older-post, where *Group* indicates age category and *State* indicates the pre- or post-standing period.

#### C. Linear mixed model

LMMs were employed to investigate which factors significantly influence the HRV-PRV consistency. Fixed effects for the LMMs were *Group* (younger vs. older), *State* (pre- vs. post-standing), *TestType* (rapid supine-to-stand, slow supine-to-stand, head-up tilting, and sit-to-stand), *Sex* (Female vs. Male), the interaction between *Sex* and *Group*, and the interaction between *State* and *Group*. Random effects were created for the subject IDs to correct the individual influence on the results. The LMM analysis was performed for each HRV-PRV pair using R Statistical Software [43]. The model is written as:

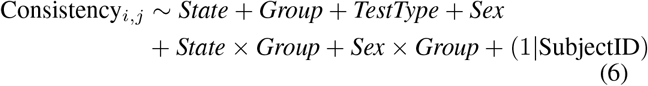

where we defined the following metric to quantify the consistency for *i*th feature:

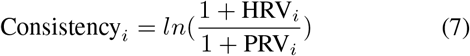

It can be observed that 0 represents PRV perfectly matches HRV. A positive consistency refers PRV underestimating the HRV and a negative consistency refers overestimation. This logarithmic transformation was primarily employed to satisfy the distributional assumptions of LMM. Since all features in this study are nonnegative, the 1+HRV and 1+PRV ensures the logarithm operation is well-defined.

To account for potential heteroscedasticity in the data, Levene’s test was first performed to assess the homogeneity of variance across different age groups and postural states. As the Levene’s tests suggested heteroscedasticity, LMMs were fitted using variance function structure (VarIdent in R) to allow for group-specific residual variances. Sequentially, Type III F-tests differentiate the impact of different fixed effects on HRV-PRV consistency, with significance level set at 0.05. Post hoc analysis was conducted using Benjamini–Hochberg procedure to control the false discovery rate (FDR), which has been used in post hoc analysis for HRV [40].

#### D. Mediation analysis

Three mediation analysis were conducted to evaluate whether mean AC, mean dP/dt_*max*_, and mean TPR (normalized by body surface area) mediate the fixed effects on HRV–PRV consistency. These hemodynamic mediators were estimated by a three-element Windkessel model using the Finapres software [45]. Due to software availability, this analysis was restricted to dataset I.

Considering the potential high collinearity, these mediators were analyzed separately for each feature. Each mediation analysis involves two LMMs: a mediator model and an outcome model. The mediator model predicts the mediator as a function of experimental factors, and the outcome model predicts the dependent variable (HRV–PRV consistency) as a function of both the mediator and the same experimental factors. The outcome model is written as:

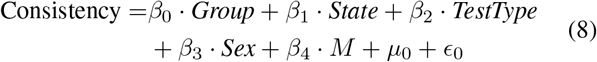

where *µ*_0_ is the random effect and *ϵ*_0_ denotes the residual. Interaction was not included in this model as it would introduce extra complexity and our focus is to investigate the separate intermediate effects of age and posture change. *β*_0_, *β*_1_, *β*_2_, *β*_3_, and *β*_4_ are regression coefficients. *M* denotes the mediators in this study. The intermediate model is:

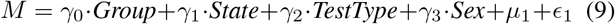

where *γ*_0_, *γ*_1_, *γ*_2_ are regression coefficients and *µ*_1_ and *ϵ*_1_ corresponds to random effect and residual.

Three metrics were utilized for mediation effect evaluation based on the introduced models. The average causal mediation effect (ACME) quantifies the indirect effect through the mediator, and the average direct effect (ADE) captures the direct effect independent of the mediator. Total effect (TE) is the summation of ACME and ADE. Eqs. 10-12 showed their definitions:

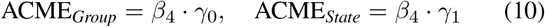

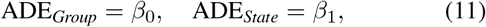

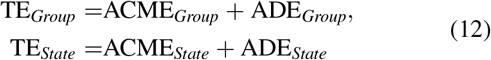

Given the residual of output models were not uniformly distributed, we employed bootstrap (5000 resamples) to estimate the CIs for each ACME, ADE and TE, which is considered as a sufficient number given the commonly chosen 2000 resamples for 90–95% CIs [44]. Mediation effects were evaluated only for the main effects *Group* and *State* as distinguishing different test types was not the primary focus of this study. The analysis was performed for each feature, resulting in 3 (mediators) × 2 (fixed effects) × 18 (features) × 3 = 324 effect estimates (including ACME, ADE, and TE). Each mediator has 108 effect estimates. To account for multiple comparisons and reduce the risk of Type I errors, we adopted 99% CIs instead of the conventional 95% CIs. This decision was based on the difficulty of deriving reliable p-values from bootstrap distributions, which may be skewed. In addition, 99% CIs implied an expected false positives number of 108 × 0.01 = 1.08 for each mediator, which is considered acceptable given the large number of comparisons.

## III. Results

### A. Data selection

Table II summarizes the number and ratio of selected segments across different test types and age groups. In total, data from 77 subjects (23 younger males, 12 younger females, 23 older males, and 19 older females) were analyzed, including 638 segments that passed the criterion. Mediation analysis was restricted to Dataset I, as only this subset provided the estimated hemodynamic variables via the Finapres Modelflow Algorithm [45]. Finally, 48 subjects (18 younger males, 7 younger females, 12 older males, and 11 older females), contributing 507 recordings, were included for mediation analysis. Fig. 3 illustrates the distribution of median and IQR of PAT across different age group, sex and postures from the 77 subjects. The median PATs stayed mostly in the range of 200ms–300ms, with an IQR of 10ms–30ms, which is similar to previous studies [26]. Fig. 4 showed the distribution of the hemodynamic variables across 48 subjects from Dataset I. It can be observed that younger adults have higher AC and lower TPR. The mean dP/dt_max_ is similar among younger and older adults.

**Table II.** Retained Data Segments Across Groups AND Protocols.

| Protocol | Group | Subject<br>[Female] | Retained/Total Segments (%) |  |
| --- | --- | --- | --- | --- |
|  |  |  | Pre-standing | Post-standing |
| Dataset I |  |  |  |  |
| Rapid supine-to-Stand | Young | 22 [6] | 52/85 (61.2%) | 53/85 (62.4%) |
|  | Old | 17 [10] | 30/64 (46.9%) | 26/64 (40.6%) |
| Slow Supine-to-Stand | Young | 15 [4] | 38/54 (70.4%) | 32/54 (59.3%) |
|  | Old | 16 [8] | 24/61 (39.3%) | 17/61 (27.9%) |
| Sit-to-Stand | Young | 23 [6] | 45/74 (60.8%) | 44/74 (59.5%) |
|  | Old | 17 [7] | 23/66 (34.8%) | 28/66 (42.4%) |
| Head-Up Tilt | Young | 8 [2] | 23/33 (69.7%) | 20/33 (60.6%) |
|  | Old | 23 [12] | 36/66 (54.5%) | 31/66 (47%) |
| Dataset II (Clinical population) |  |  |  |  |
| Rapid supine-to-stand | Old | 17 [6] | 38/72 (52.8%) | 21/72 (29.2%) |
| Dataset III |  |  |  |  |
| Rapid supine-to-stand | Young | 10 [5] | 29/29 (100%) | 27/29 (93.1%) |
| Dataset I subset for mediation analysis (after ABP selection) |  |  |  |  |
| Rapid supine-to-Stand | Young | 22 [6] | 49/85 (57.6%) | 52/85 (61.2%) |
|  | Old | 16 [9] | 27/64 (46.9%) | 26/64 (40.6%) |
| Slow Supine-to-Stand | Young | 15 [4] | 37/54 (68.5%) | 32/54 (59.3%) |
|  | Old | 15 [7] | 24/61 (39.3%) | 16/61 (26.2%) |
| Sit-to-Stand | Young | 23 [6] | 45/74 (60.8%) | 44/74 (59.5%) |
|  | Old | 17 [7] | 22/66 (48.5%) | 28/66 (45.5%) |
| Head-Up Tilt | Young | 8 [2] | 23/33 (69.7%) | 20/33 (60.6%) |
|  | Old | 21 [10] | 32/66 (48.5%) | 30/66 (45.5%) |

**Fig. 3.**
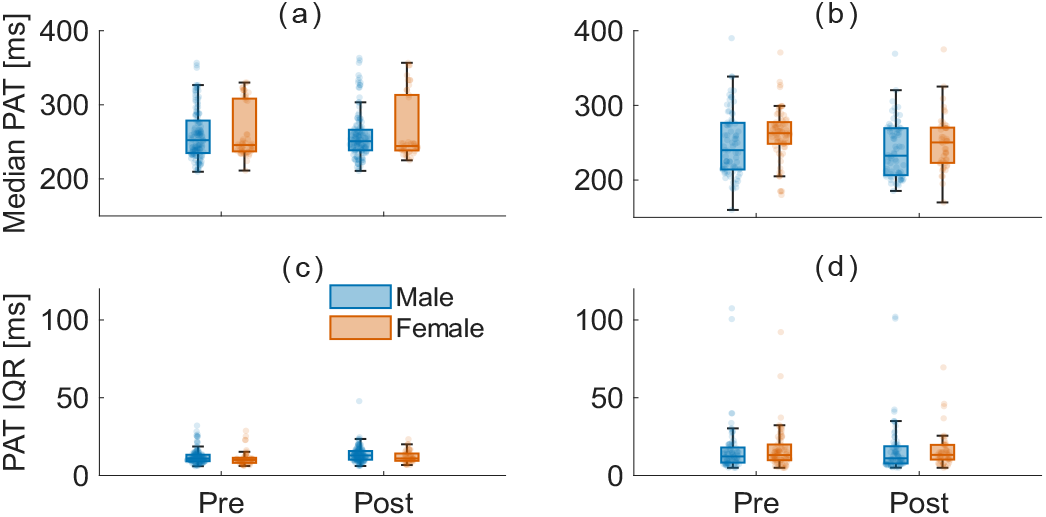
Boxplot of the median and interquartile range (IQR) of PAT across subgroups. Tukey = 1.5*(Q3-Q1). Blue denotes male subjects, orange denotes female subjects. (a) Median PAT of younger adults; (b) Median PAT of older adults; (c) PAT IQR of younger adults; (d) PAT IQR of older adults.

**Fig. 4.**
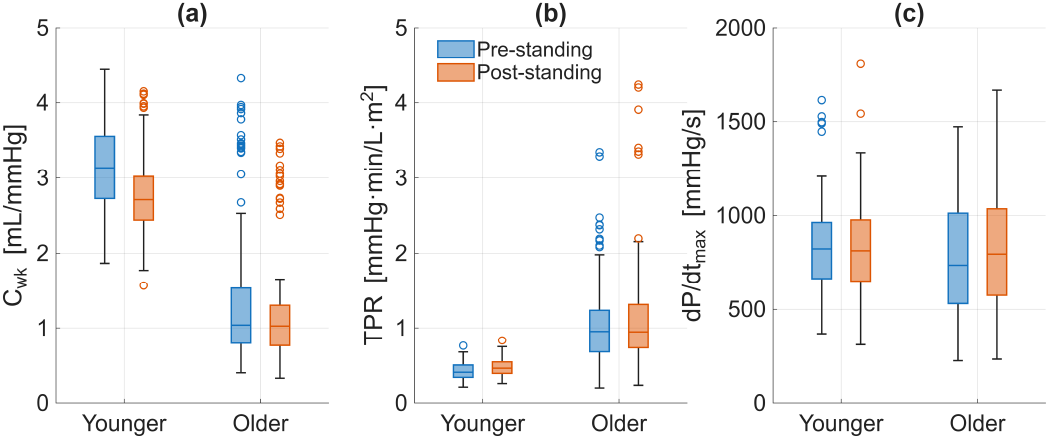
Hemodynamic baseline characteristics across subgroups and postures. (a) Mean arterial compliance, (B) Mean total peripheral resistance, and (C) Mean dP/dt_max_ (cardiac contractility) categorized by age group (Younger vs. Older) and postural state (Pre- vs. Post-standing). Tukey = 1.5*(Q3-Q1).

#### B. Bland-Altman analysis

Fig. 5 and 6 summarize the results of Bland–Altman analysis. Each subfigure displays five bars, representing the bias and LoA width estimates for the entire sample and the four subgroups. A limited number of features’ biases had 95% CIs including zero: AVNN post-standing in both age groups; SD2 for older adults pre-standing, ApEn, alpha1 and alpha2 for younger adults pre-standing, and alpha2 for older adults post-standing. Overall, most features did not demonstrate population-level HRV-PRV consistency.

**Fig. 5.**
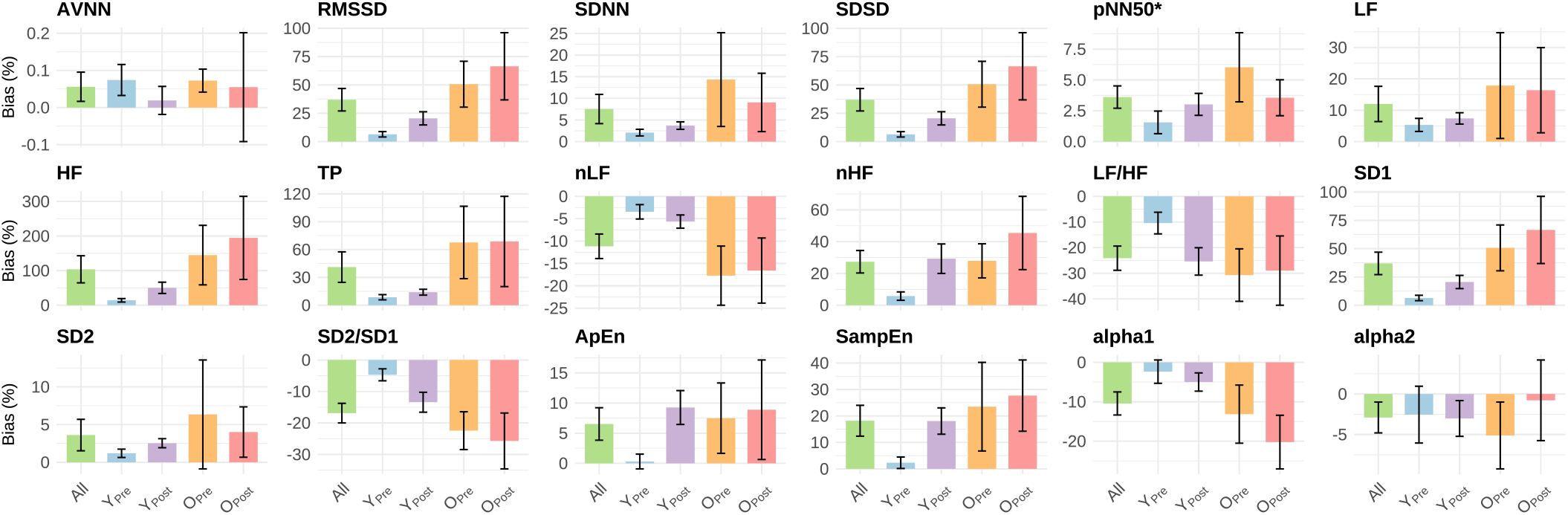
Biases with their 95% CIs for each feature in the entire population and in different subgroups. All results were normalized by HRV except for pNN50. All: results fromentire population Y: younger adults; O: older adults; Pre: Pre-standing; Post: Post-standing. Asterisk denotes absolute bias instead of normalized bias.

**Fig. 6.**
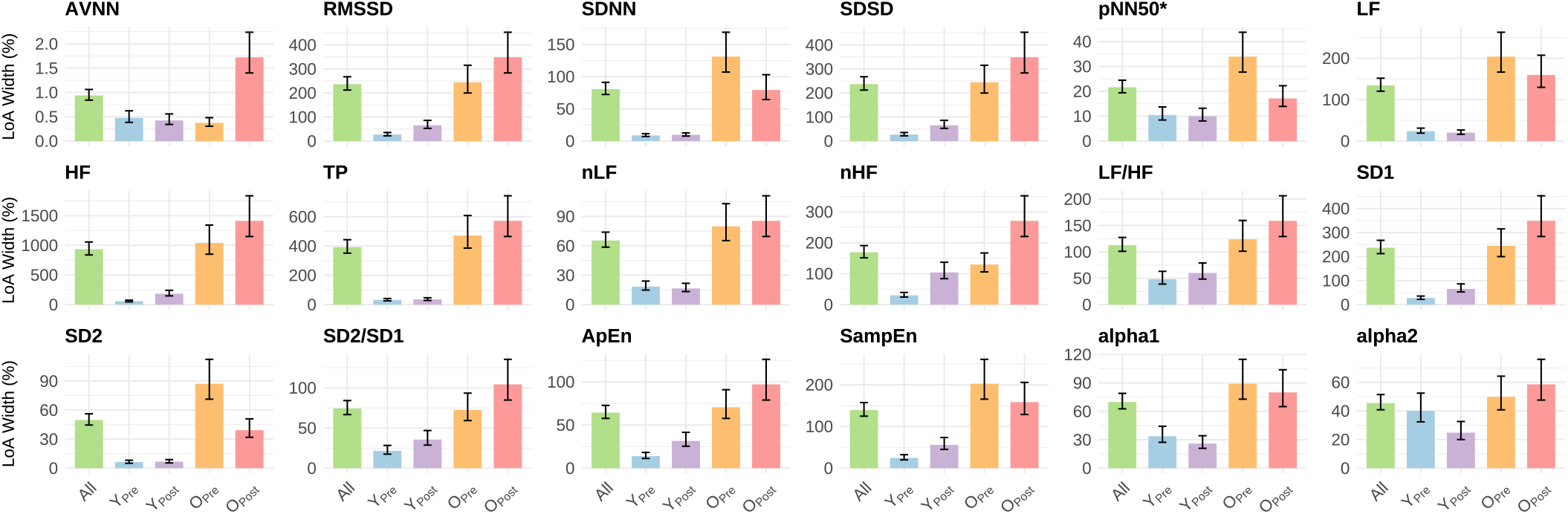
LoA width with their 95% CIs (error bars) for each feature in the entire population and in different subgroups. All results were normalized by HRV except for pNN50. All: results fromentire population. Y: younger adults; O: older adults; Pre: Pre-standing; Post: Post-standing. Asterisk denotes absolute bias instead of normalized bias.

At the individual-level, all features from younger adults met the 150% LoA width threshold except for post-standing for HF. Conversely, individual-level agreement was markedly reduced in older adults, where only AVNN, nLF, SD2, ApEn, SD2/SD1, alpha1, and alpha2 remained within the 150% boundary. Additionally, across the majority of features, younger adults exhibited significantly narrower LoA widths compared to older adults except for AVNN and alpha2. These results highlighted the fundamental age-related decline in HRV-PRV agreement.

#### C. Linear mixed model

Table III presents the *p*-values derived from type III F-tests after FDR correction. Both aging and posture change have a significant effect on the HRV-PRV agreement: the effect of age (*Group*) was significant for 12 features; postural change effect (*State*) showed significance for 10 features. *TestType* also showed significance for 10 features. While the main effect of *Sex* was significant for only three features, interaction effects between *Sex* and *Group* was significant for six features. The age (*Group*) and posture (*State*) interaction was significant for entropy measures, pNN50, and LF/HF. The interaction plots these effects are provided in Supplementary Material II to visualize the difference in agreement across subgroups.

**Table III.** P-values derived from type III F-tests after Benjamini–Hochberg process (FDR correction). P-values < 0.05 were highlighted.

| Feature | Age<br>(*Group*) | Posture<br>(*State*) | Test<br>type | Sex | Age ×<br>Posture | Age ×<br>Sex |
| --- | --- | --- | --- | --- | --- | --- |
| AVNN | 0.49 | **0.02** | **<0.01** | 0.20 | 0.79 | 0.09 |
| RMSSD | **<0.01** | **0.04** | **0.02** | 0.16 | 0.41 | 0.051 |
| SDNN | **0.04** | 0.55 | **0.01** | **0.03** | 0.79 | **0.02** |
| SDSD | **<0.01** | **0.04** | **0.04** | 0.25 | 0.40 | 0.09 |
| pNN50 | **<0.01** | 0.28 | 0.13 | **0.03** | **0.04** | **<0.01** |
| LF | 0.63 | 0.20 | 0.06 | 0.26 | 0.40 | **0.03** |
| HF | **0.01** | **0.04** | **<0.01** | 0.55 | 0.38 | 0.16 |
| TP | **<0.01** | 0.39 | **0.01** | 0.16 | 0.82 | **0.03** |
| nLF | **<0.01** | 0.52 | 0.12 | 0.51 | 0.42 | 0.48 |
| nHF | **<0.01** | 0.19 | **0.02** | 0.87 | 0.26 | 0.59 |
| LF/HF | 0.12 | **<0.01** | **<0.01** | 0.57 | **0.03** | 0.72 |
| SD1 | **<0.01** | **0.04** | **0.02** | 0.16 | 0.40 | 0.051 |
| SD2 | 0.28 | 0.27 | **0.02** | 0.07 | 0.95 | **0.049** |
| SD2/SD1 | **<0.01** | **0.01** | 0.15 | 0.42 | 0.37 | 0.17 |
| ApEn | 0.83 | **<0.01** | 0.87 | 0.42 | **<0.01** | 0.25 |
| SampEn | **<0.01** | **<0.01** | 0.45 | 0.48 | **<0.01** | 0.58 |
| alpha1 | **<0.01** | **0.02** | 0.17 | 0.78 | 0.73 | 0.42 |
| alpha2 | 0.27 | 0.54 | 0.55 | **<0.01** | 0.49 | **0.02** |

#### D. Mediation analysis

Fig. 7–9 illustrate the results of the mediation analysis. A mediation effect was considered significant if the 99% CI excluded zero, with significance indicated by asterisks.

**Fig. 7.**
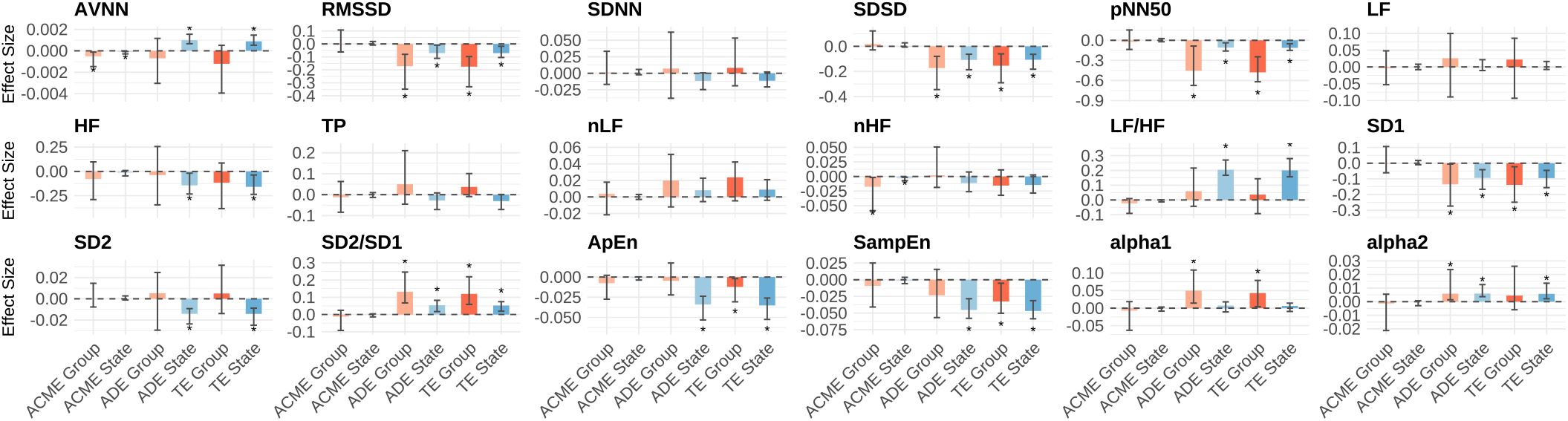
ACME, ADE and TE in different features for mean arterial compliance. For each feature, six bars represent the estimated effect sizes for variable *Group* (in red) and *State* (in blue). Error bars indicate 99% CIs. Asterisks denote statistically significant effects, defined as CIs that do not cross zero. ACME_*State*_, ADE_*State*_, etc. represent the group- and state-level mediation effects as defined in Eq. 10-12.

**Fig. 8.**
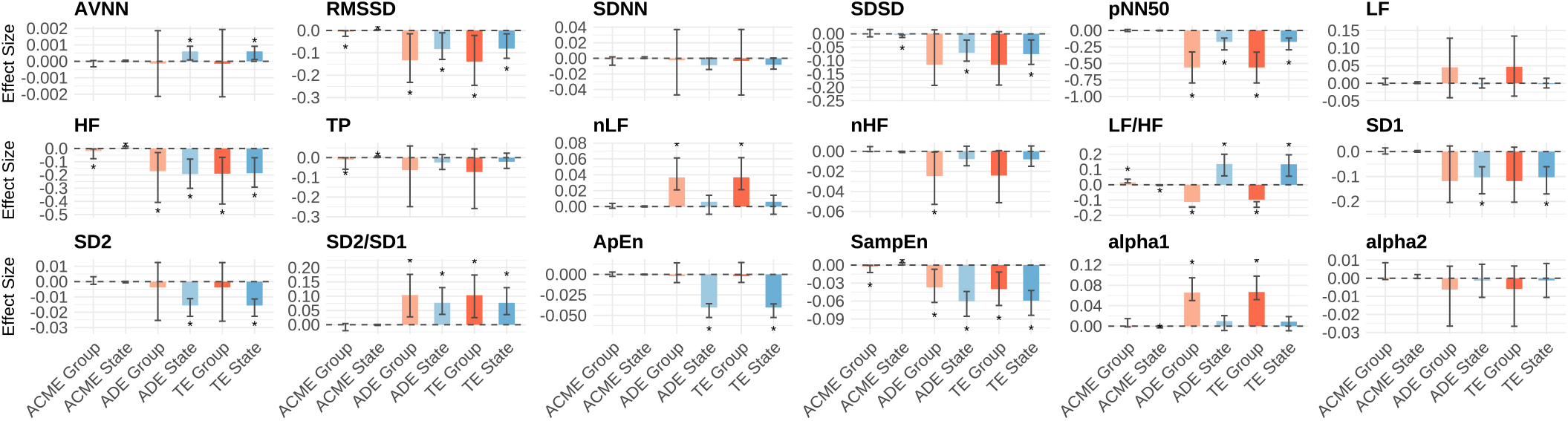
ACME, ADE and TE in different features for mean dP/dt_max_ (cardiac contractility). For each feature, six bars represent the estimated effect sizes for variable *Group* (in red) and *State* (in blue). Error bars indicate 99% CIs. Asterisks denote statistically significant effects.

**Fig. 9.**
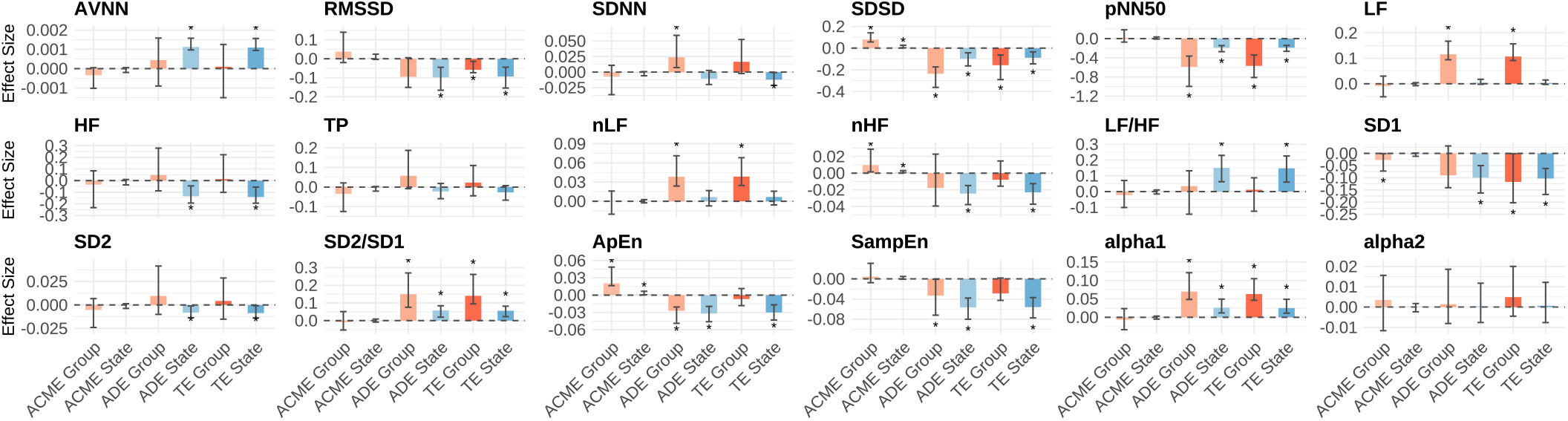
ACME, ADE and TE in different features for mean total peripheral resistance. For each feature, six bars represent the estimated effect sizes for variable *Group* (in red) and *State* (in blue). Error bars indicate 99% CIs. Asterisks denote statistically significant effects.

Among all three mediators, ADEs were consistently larger than ACME in terms of both significance and effect size. Specifically, for posture (*State*), ADE reached significance for 12, 11, and 13 features in the AC, dP/dt_max_, and TPR models, respectively. For the impact of Age (*Group*), significant ADEs were observed for seven (AC), nine (dP/dt_max_), and nine (TPR) features.

Despite the predominance of direct effects, a few features exhibited limited but statistically significant mediation effects. For mean AC, both AVNN and nHF showed significant ACME_*Group*_ and ACME_*State*_. For dP/dt_max_, RMSSD, HF, TP, LF/HF, and SampEn, showed significant but small ACMEs for both age and posture change. Additionally, SDSD and alpha1 exhibited significant ACME_*State*_. For mean TPR, significant age and posture mediation were identified for SDSD, nHF, and ApEn. Notably, we observed a inconsistent mediation effects for SDSD and ApEn, where the sign of the ACME was opposite to the ADE.

## IV. Discussion

This study systematically evaluated HRV–PRV interchangeability in younger and older adults across multiple postural changes, and examined the potential mediating roles of mean arterial compliance, cardiac contractility, and TPR. The independent impacts of aging, posture change, and sex, as well as the interactions between aging, sex, and postural change, were analyzed using LMM. We conclude that the agreement is generally higher in younger adults, with many features are interchangeable. In contrast, in older adults, most features indicating fast physiological regulation dynamics are non-interchangeable, although some slower indices remained interchangeable. Both aging and postural change independently reduced HRV–PRV consistency. Males and females exhibited distinct patterns of decline, especially in slower metrics such as SDNN and alpha2. Vascular and hemodynamic measures were not sufficient to explain the HRV–PRV agreement decline caused by aging and postural change.

### A. Limited interchangeability of HRV and PRV

Bland–Altman analysis demonstrated that interchangeability was higher in younger adults than in older adults. This difference was most evident at the individual level, where all indices except HF were interchangeable in younger adults. The reduced agreement in older adults suggests that PRV should be interpreted with caution in this population. This pattern likely reflects that vascular aging affected all features but have a larger effect on the interchangeability of faster indices. Across all features, the LoA widths for younger adults remained significantly narrower than older adults, underscoring a fundamental decline in PRV reliability with aging. Importantly, the direction of bias was consistent across subgroups, supporting the deviations are unlikely to be explained solely by random errors.

Frequency-domain indices (HF and TP) exhibited the poorest consistency, as evidenced by large LoA widths. It is recognized that the arterial vessels typically act as a low-pass filter for pulse wave transmission [20], and vascular aging modifies this filtering transfer function. Vascular aging (AC decrease) therefore might result in larger PTT oscillations, causing PRV to deviate significantly from HRV, particularly in HF and TP. Notably, HF also showed to exceed the threshold for younger adults post-standing, suggesting that the HRV-PRV relationship was largely impaired after posture change. In contrast, nonlinear features demonstrated relatively narrower LoA widths compared to frequency-domain measures, indicating they may be more robust to the vascular impact introduced by pulse wave.

It should be noticed that we took the average across different test types for each subject within each state (pre- or post-standing) to ensure statistical independence. Nevertheless, it inevitably collapses the maneuver variability among different orthostatic challenges. Given that the LMM identified *TestType* as a significant effects for many features, this averaging may lead to wider LoA, as the variance between different physiological stressors is incorporated into the overall error estimate. Consequently, the reported LoA width should be interpreted as a conservative estimation of PRV-HRV agreement for its deployment in individuals.

Taken together, these findings indicate that PRV can serve as a substitute for HRV in younger healthy adults when appropriate bias correction is applied. In older adults, however, HRV and PRV appear largely non-interchangeable, suggesting that PRV should be regarded as a distinct digital biomarker rather than a direct surrogate for HRV.

### B. Impact of aging

LMM analysis suggested aging independently reduced HRV–PRV consistency across 12 features. This aligns with previous findings that HRV–PRV interchangeability is lower in clinical populations with cardiopulmonary and neurological diseases [20]. These results also support the idea that HRV–PRV consistency itself could serve as a novel digital biomarker for vascular and autonomic health, warranting further validation in daily monitoring settings.

In addition, we found that features presenting longer-scale variability mostly exhibited significance for age×sex interaction instead of just age, e.g., SDNN, pNN50, LF, TP, and alpha2. Furthermore, RMSSD exhibited a borderline significant effect after FDR correction (*p* = 0.051). These results suggest that HRV–PRV agreement decline may be a sex-specific manifestation of vascular aging. Given the widespread commercial use of SDNN and RMSSD, wearable diagnostics must adopt gender-aware models in elderly populations.

Despite the clear impact of aging, our mediation analysis revealed that the age-related decline in HRV–PRV consistency only showed minor mediation effects by mean AC, cardiac contractility, or TPR. While it is regarded pulse transition time as primary source of HRV-PRV inconsistency, and is linked to TPR and AC, we suggest that the HRV-PRV discrepancies likely reflects a robust physiological decoupling that captures unique cardiovascular information.

### C. Impact of postural change

We found postural change also greatly decrease HRV-PRV consistency [18], [23], despite some prior studies suggested PRV and HRV have good agreement [19], [21], [22]. Compared with the previous studies, we included more subjects (35 younger and 42 older adults), including multiple types of postural changes and repeated sessions, which should improve generalizability of the results. We found that posture change induced a pronounced reduction in HRV-PRV consistency for the entropy measures. Moreover, significant age×posture interaction effects were also identified. This is evidenced by the Bland-Altman analysis: while younger adults exhibited a marked increase in systematic bias following postural challenges, the bias in older adults remained relatively stable. Future work should further investigate the physiological interpretation behind the distinct entropy consistency change across different population.

It should be noted that the variable *TestType* exhibited a significant effect across multiple features, suggesting that the degree of HRV-PRV consistency is sensitive to different types of postural change. It is physiologically plausible that more vigorous maneuvers, such as HUT or supine-to-stand, elicit more profound orthostatic stress and autonomic activation compared to a simple sit-to-stand transition. This distinction might be critical for standardizing clinical protocols and defining the operational boundaries of PRV as a surrogate for HRV in dynamic or ambulatory environments.

Similarly to aging, while postural changes significantly disrupted the HRV–PRV agreement, this effect cannot be primarily explained by the mean AC, TPR, or cardiac contractility change. The fundamental physiological reasons behind the observed HRV-PRV consistency decline post posture change still needs to be further addressed.

### D. Impact of sex

This study also revealed a sexual dimorphism in the HRV-PRV agreement. Specifically, the consistency of SDNN, pNN50 and alpha2 was governed by both the significant sex, and age-sex interaction effects. Notably, while the main effect of age on LF and alpha2 was less pronounced, the interaction effects underscore that the aging trajectory of HRV-PRV consistency is fundamentally different for men and women. Voss et al. reported that ECG derived alpha2 follows a relatively gender-independent trajectory during aging [46]. Therefore, our observed sex-specific significance is likely driven by the divergent vascular aging patterns, specifically the accelerated arterial stiffening and endothelial dysfunction experienced by post-menopausal females [47], [48].

For LF, TP, and SD2, the Age–Sex interaction reached statistical significance (*p* < 0.05) when the main effect of *Sex* remained non-significant. This emphasizes that the impact of sex on HRV–PRV consistency might be a dynamic process that intensifies with aging. These significant interaction effects suggest that the transition from young to old might induce more severe peripheral circulation in females than in males.

### E. Clinical and Engineering implications

Our results highlight that PRV may not be a reliable substitute for HRV especially in older adults, which has direct consequences for its use in wearable devices and remote monitoring systems. Although reference values for HRV during daily activities in older adults have been reported [49], [50], these reference values cannot be directly transferred to PRV. HRV and PRV should be explicitly distinguished in both clinical interpretation and wearable-based assessment. In addition, our findings suggest that wearable algorithms should incorporate age-, posture- and sex-aware calibration strategies. Given the significant interaction effects of age and sex observed in this study, such calibration is important as vascular aging progresses differently in males and females. Without this consideration, PRV-derived indices may be misinterpreted as reflecting autonomic dysfunction when they are, at least in part, influenced by vascular and hemodynamic factors. Furthermore, because traditional hemodynamic mediators such as AC and TPR did not fully account for the decline in HRV– PRV agreement, the discrepancy between HRV and PRV may itself provide useful physiological information. Rather than being treated solely as measurement error, this divergence may reflect more complex impairment in cardiovascular regulation and could serve as a potential marker for vascular health monitoring.

### F. Limitations

This study has some limitations. First, the duration used for HRV and PRV computation was relatively short, and the window length for pre-standing and post-standing is unequal. Window length shorter than five minutes is commonly referred as ultra-short-term HRV [2]. Although it may raise concerns regarding the stability of feature estimation, previous research has demonstrated that a three-minute and five-minute recording provided similar HRV-PRV agreement level for time, frequency and a few nonlinear features [51]. However, alpha2 is not included and should thus be interpreted with caution. In addition, despite strict criteria were utilized, feature values and conclusions could still be influenced by subtle peak detection inaccuracies. Second, although significant sexual difference in HRV–PRV agreement was identified, the relatively small sample size of the younger female subgroup limits our ability to define definitive, sex-specific interchangeability thresholds; thus, larger balanced cohorts are recommended for further subgroup-specific validation. Finally, this study categorizes subjects into younger and older groups to facilitate subgroup comparisons. Future studies with larger, more balanced cohorts are required to establish more precise age-specific interchangeability thresholds.

## V. Conclusion

This study examined the interchangeability between HRV and PRV in young and older adults across multiple postural changes. We concluded that the ultra-short features studied in this study are mostly interchangeable for younger adults even after posture change. For older adults, they are largely non-interchangeable. Both aging and postural change reduced HRV–PRV agreement. Furthermore, we observed that males and females exhibited distinct patterns in the HRV–PRV agreement decline. These findings underscore the need for cautious interpretation when relying on PRV in daily monitoring and provide physiological insight into the mechanisms underlying HRV–PRV non-interchangeability. Moreover, the observed discrepancies suggest that HRV–PRV inconsistency holds potential as a novel biomarker for autonomic cardiovascular health monitoring.

## Data Availability

All data produced are available online at：https://doi.org/10.34973/te70-x603

https://data.ru.nl/collections/di/dcn/DSC_626810_0007_173

## Acknowledgment

The authors would like to acknowledge the help from Dr. Jurgen A.H.R. Claassen, Dr. Arjen Mol, and Annemijn Hoff on this study. This study is funded in part by China Scholarship Council.

## Notes

### Competing Interest Statement

The authors have declared no competing interest.

### Funding Statement

This study did not receive any funding

### Author Declarations

Ethics Committee of Radboud University Medical Center gave ethical approval for the original study (https://doi.org/10.34973/te70-x603). The present work is a secondary analysis of this dataset, which is available through the Donders Institute Research Data Repository under a Data Use Agreement. No new data were collected.

### Summary of Updates

1. New datasets included 2. Bland-Altman analysis modified 3. LMM mediators modified 4. Adding sex - in LMM

