## Supplementary Material for "Aging and Postural Changes Influence the Agreement Between Pulse Rate Variability and Heart Rate Variability"

### Supplementary Material I: Summary of HRV, PRV, PATV and mean BP calculation

Fig. 1 presents the scatter plots between different HRV and PRV features with the identity line, among different age group, sex and postures. Different test types were not differentiated given the page limitation. For LF, HF, LF/HF and SD2/SD1, values were log-10 transformed for better visualization. Dashed line represents the identity line (HRV=PRV). Blue points (younger adults) consistently align with the identity line, indicating high PRV-HRV agreement. In contrast, red points (older adults) exhibit increased dispersion, highlighting the degradation of PRV reliability. Several indices reflecting long-term variability (AVNN, SDNN, LF, and SD2) exhibited better alignment with the identity line across both younger and older adults. Other showed notably greater dispersion, particularly in the elderly cohort.

### Supplementary Material II: Interaction effects

Fig. 2 shows the interaction plots (age $\times$posture) for each feature. Each point represents the average model-predicted response *State* × *Group*, after marginalizing over the fixed-effect covariates and excluding the random effects and residual variance. Error bars depict 95% CIs derived from fixed-effects standard errors. According to our defined consistency metric, consistency closer to zero indicates better agreement. It can be observed that both age groups showed similar changing patterns during tests across most features, except ApEn.

Fig. 3 shows the age-sex interaction plots for each feature. It revealed that the impact of aging on HRV-PRV agreement is modulated by sex. Specifically, a more significant reduction in HRV-PRV consistency (such as SDNN, pNN50, SD1, and alpha2) was found in older females compared to older males, whereas these sex differences were not significant in the younger group. This interaction potentially reflects the sex-specific vascular aging and autonomic regulation changes after menopause.

Runwei Lin and Ying Wang are with the Department of Biomedical Signals and Systems, University of Twente, Enschede, the Netherlands.

Marjolein Klop is with the Department of Geriatric Medicine, Radboud University Medical Center, Nijmegen, the Netherlands

Richard van Wezel is with the Department of Neurobiology, Donders Institute for Brain, Cognition and Behaviour, Radboud University, Nijmegen, the Netherlands, and with the Department of Biomedical Signals and Systems, Technical Medical Centre, University of Twente, Enschede, the Netherlands, and also the OnePlanet Research Center, Radboud University, Nijmegen, the Netherlands.

Frank R. Halfwerk is with the Cardiac Surgery Innovations Lab, Engineering Organ Support Technologies Group, Department of Biomechanical Engineering, University of Twente, Enschede, the Netherlands, and with the Department of Cardio-thoracic surgery, Thorax Centrum Twente, Medisch Spectrum Twente, Enschede, the Netherlands, and with the Cardiovascular Health Technology Centre, TechMed Centre, University of Twente, the Netherlands.

Dirk W. Donker is with the group of Cardiovascular and Respiratory Physiology, University of Twente, Enschede, the Netherlands, and with Intensive Care Center, University Medical Center Utrecht, Utrecht, the Netherlands.

Gozewijn Dirk Laverman is with the Department of Biomedical Signals and Systems, University of Twente, and with the Department of Internal Medicine, Ziekenhuisgroep Twente, Almelo, the Netherlands.

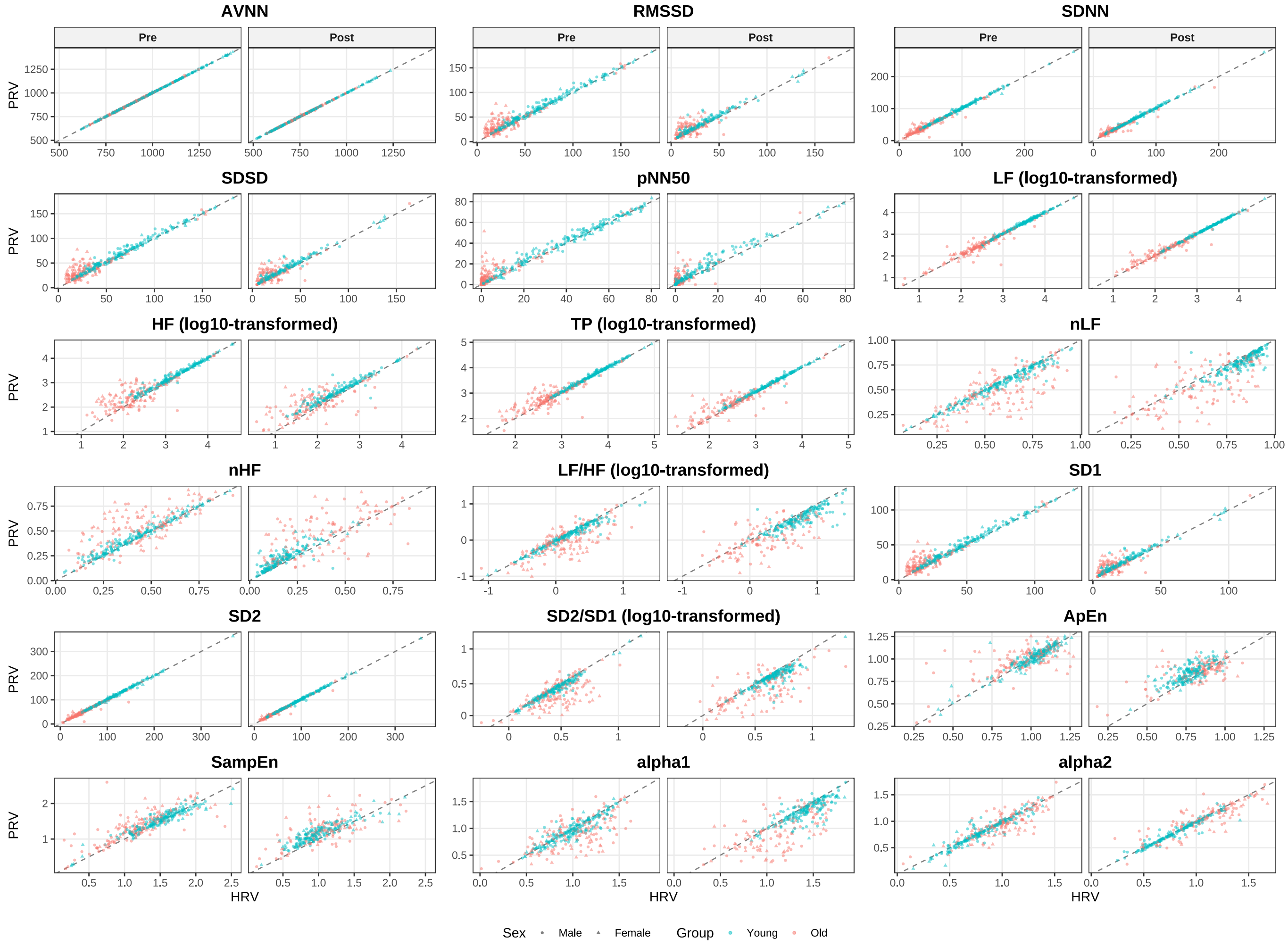


Fig. 1. Scatter plots of 18 HRV and PRV features with identity line (dashed line), X axis is HRV value and y axis is PRV. Each subplot compares the HRV and PRV during Pre-standing (left) and Post-standing (right) phase. The value of LF, HF, TP, LF/HF and SD2/SD1 were $log10$ transformed for better visualization. Blue points denote younger adults, and red points were older adults. Rectangular points denote male and triangles denote female.

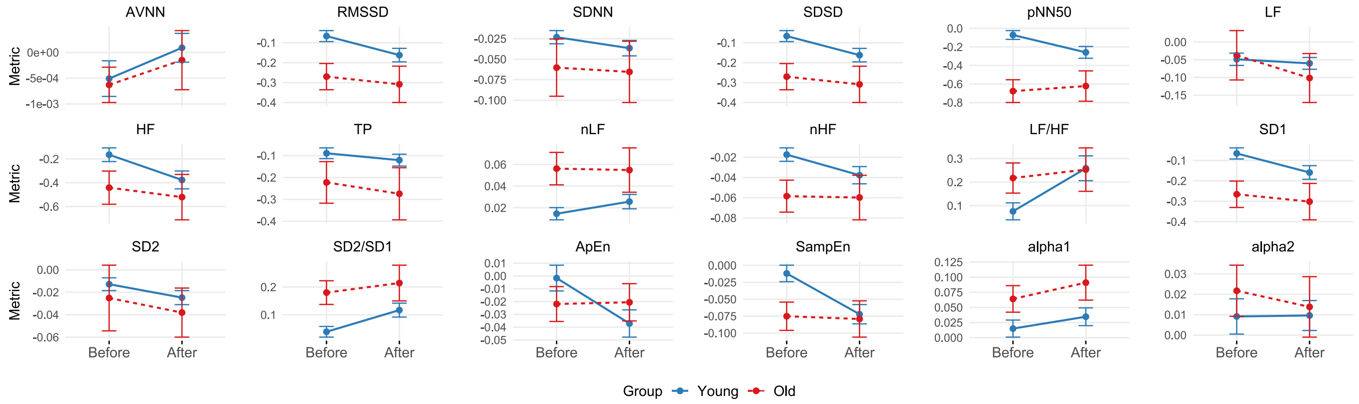


Fig. 2. Interaction plots for HRV-PRV consistency change in different postural states and age groups after marginalizing other effects. Red lines denote older adults and blue line denote younger adults.

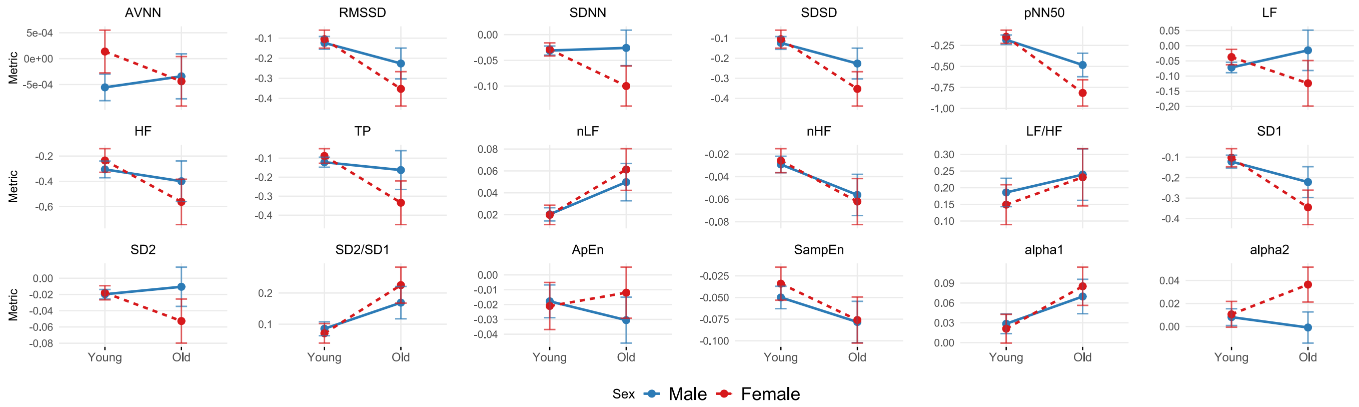


Fig. 3. Interaction plots for HRV-PRV consistency change in different age and sex groups after marginalizing other effects. Red lines denote female and blue lines denote males.
